# Evaluating the Portability of SepsisWatch: A Multi-Site External Validation of a Sepsis Machine Learning Model

**DOI:** 10.1101/2024.03.22.24304753

**Authors:** Bruno Valan, Anusha Prakash, William Ratliff, Michael Gao, Srikanth Muthya, Ajit Thomas, Jennifer L. Eaton, Matt Gardner, Marshall Nichols, Mike Revoir, Dustin Tart, Cara O’Brien, Manesh Patel, Suresh Balu, Mark Sendak

## Abstract

**Importance:** Sepsis accounts for a substantial portion of global deaths and healthcare costs. Early detection using machine learning (ML) models offers a critical opportunity to improve care and reduce the burden of sepsis.

**Objective:** To externally validate the SepsisWatch ML model, initially developed at Duke University, in a community healthcare and assess its performance and clinical utility in early sepsis detection.

**Design:** This retrospective external validation study evaluated the performance of the SepsisWatch model in a new environment. Data from patient encounters at Summa Health’s emergency departments between 2020 and 2021 were used. The study analyzed the model’s ability to predict sepsis using a combination of static and dynamic patient data.

**Setting:** The study was conducted at Summa Health, a nonprofit healthcare system in Northeast Ohio, covering two emergency departments (EDs) associated with acute care hospitals, and two standalone EDs.

**Participants:** Encounters associated with adult patients in any of Summa Health’s four EDs were included. Encounters lasting <1 hour were excluded. Only the first 36 hours of each encounter were used in model evaluation.

**Intervention(s)/Exposure(s):** The SepsisWatch model was used to predict sepsis based on patient data.

**Main Outcome(s) and Measure(s):** The primary outcomes measured were the model’s area under the precision-recall curve (AUPRC), and area under the receiver operator curve (AUROC).

**Results:** The study included 205,005 encounters from 101,584 unique patients. 54.7% (n = 112,223) patients were female and the mean age was 50 (IQR, [38,71]). The model demonstrated strong performance across the Summa Health system, with little variation across different sites. The AUROC ranged from 0.906 to 0.960, and the AUPRC ranged from 0.177 to 0.252 across the four sites.

**Conclusions and Relevance:** The external validation of the SepsisWatch model in a community health system setting confirmed its robust performance and portability across different geographical and demographic contexts. The study underscores the potential of advanced ML models in improving sepsis detection in both academic and community hospital settings, paving the way for prospective studies to measure the clinical and operational impact of such models in healthcare.

## Introduction

Sepsis is a systemic inflammatory response to disseminated infection that affects millions of people worldwide.^1^ Despite medical advancements, sepsis continues to be a major health concern, accounting for 19.7% of all global deaths.^2^ In the US, sepsis is the cause of up to half of all in-hospital fatalities.^3^ The economic impact of sepsis is equally staggering. Between 2012 and 2018, the annual cost of inpatient hospitalizations related to sepsis for Medicare beneficiaries increased 26.12% from $17.8 billion to $22.4 billion.^4^ Costs for each sepsis-related encounter were estimated to be $18,023 when identified before admission and $51,022 if recognized post-admission.^5^ Importantly, early detection and prompt antibiotic treatment can significantly reduce both the cost and the health impacts of sepsis.^6–11^ Sepsis care bundles have been developed and diffused to advance the standard of care for sepsis management, but the outcomes of these interventions are mixed.^12–14^ Leveraging machine learning (ML) for early sepsis identification has become a pivotal opportunity to further improve sepsis care.

ML models can effectively identify sepsis early and implementation of these models can influence treatment and outcomes.^15–18^ Prospective implementation of ML and non-ML driven sepsis alert systems have been shown to decrease in-hospital mortality, organ failure, and length of stay.^19–22,24^ However, other studies demonstrate that while these systems may be able to predict impending sepsis they can fall short of changing care delivery and patient outcomes.^23^ The role of ML in sepsis treatment merits continued study of models and implementation approaches in different healthcare settings and patient populations.

A key challenge to scaling ML models in healthcare is lack of portability, in part due to significant variability of patient populations and care delivery processes across geographies and points in time. Performance of ML models across diverse healthcare settings and patient populations is highly variable.^24–27^ Model deployment itself brings forth its own set of challenges. From data curation to quality assurance and continuous monitoring, successful integration requires a multifaceted and deliberate approach.^28,29^ In addition, designing and implementing a workflow solution for clinicians to act on the outputs of an ML model requires additional training and personnel.^30^ Clinical perceptions of ML interventions will also vary depending on intuition and understanding, demonstrating that model accuracy does not necessarily equate to trust by providers and overall solution effectiveness.^31^ Taken together with the privacy concerns related to use and sharing of protected healthcare data, external validations of ML models are limited.

In this paper, we describe the external validation of the SepsisWatch model in Summa Health, a hospital system in Ohio—a setting both geographically and temporally distinct from the context of initial development at Duke University. SepsisWatch is the first deep learning model implemented in routine clinical care in the United States and has been in continuous operations at Duke University since November 2018. This study has two primary objectives. First, we aim to assess the model’s performance in this new environment. Second, we aim to estimate the potential workload and benefit to patients associated with the prospective implementation of SepsisWatch in this new environment. Through this study, we hope to contribute meaningful insights into the broader applicability of ML models for sepsis detection, emphasizing the importance of context and adaptation.

## Methods

### Setting

Summa Health, a nonprofit integrated healthcare delivery system in Northeast Ohio, encompasses two acute care hospitals: Summa Health Akron City Hospital (ACH) and Summa Health Barberton (SHB), each equipped with their own emergency department (ED). The system is further complemented by two standalone EDs—ACH Green ED and SHB Wadsworth ED. These facilities form a 1,300-bed system that facilitates over one million patient encounters annually. Summa Health primarily serves Summit, Wayne, Medina, Portage and Stark Counties in Ohio. The service area encompasses urban, suburban, and rural areas. The payer mix of Summa Health patients is 5% uninsured, 25% privately insured, 30% Medicaid, and 40% Medicare.

### Data

SepsisWatch was externally evaluated on encounters for adult patients (age >= 18) who presented to one of Summa Health’s four emergency departments (EDs) between 1/1/2020 and 12/31/2021. Encounters started at the time of presentation to the ED and ended at time of discharge or death. Encounters were attributed to the site of origination, such that an encounter that began at a free-standing ED and included a transfer to an acute hospital was assigned to the free-standing ED. Each individual visit to the emergency department was considered as a separate encounter, regardless of the number of visits made by the same patient. Encounters with a length of stay less than 1-hour were excluded from the cohort and only the first 36 hours of each encounter were used in the model evaluation.

The model combines static data that remains unchanged throughout an encounter and dynamic data that is updated during an encounter. Static variables included patient demographics, encounter details, and comorbidity data, which looked at ICD-10 codes documented at any encounter in the 12 months prior to ED presentation. Dynamic data used by the model includes analyte results, vital signs, and medication administrations. Dynamic variables were considered between the encounter start time and end time.

### Outcome Definition

We used a previously developed sepsis phenotype as our outcome label. Specifically, we defined sepsis as the co-occurrence of all 3 following criteria: 1) At least two Systemic Inflammatory Response Syndrome (SIRS) criteria, which is valid for 24 hours and includes temperature anomalies (>100.4 F or <96.8 F), a heart rate above 90, a respiration rate exceeding 20, and an abnormal white blood cell count (>12 or <4); (2) a blood culture order; (3) indication of any end-organ damage, characterized by elevated creatinine (>2.0), INR (>1.5), total bilirubin (>2.0), decreased platelet count (<100), lactate levels of 2 or higher, or systolic blood pressure below 90 mmHg or a drop of 40 mmHg in systolic blood pressure within 6 hours. The varying sampling rates for medical measurements were accounted for by adjusting the relevant time window for each criterion. Vital sign documentation values (temperature, heart rate, respiration rate, blood pressure) were valid for a six-hour time window, whereas analyte measurements (white blood cell count, creatinine, bilirubin, platelet count, lactate) and orders (blood culture) were valid for a 24-hour time window. In addition, patients who met the criteria for sepsis within 1 hour of presentation to the ED were excluded.

SepsisWatch was evaluated using a detection window of 12-hours, meaning once a prediction breached the threshold, the prediction would only be classified as a true positive if the patient met sepsis criteria within 12 hours. If the prediction breached the threshold more than 12 hours prior to sepsis, the prediction was classified as a false positive. SepsisWatch was ran hourly on the hour, and produced predictions for all encounters between presenting to the ED and the minimum of time of sepsis, time of death, time of discharge, and 36 hours after ED presentation. Once a prediction breached the threshold, predictions over the next 8 hours were suppressed or snoozed. This 8-hour snooze window was designed to reduce false positive alerts and downstream potential alert fatigue. The snooze window also avoided inflating performance metrics by repetitively counting true positives.

### Model

The original model was designed at Duke University Hospital using EHR encounter data from October 1^st^ 2014 – December 1^st^ 2015. It is a recurrent neural network (RNN) model and is hereby referred to as SepsisWatch. The data used to train the model was split 80:10:10 for training, internal validation and testing, respectively. The original model performed well in multiple settings. Specifically, on an internal validation cohort SepsisWatch achieved an area under the receiver operator curve (AUROC) of 0.882 and on a temporal validation cohort the model achieved an AUROC of 0.943. The full details of the model development and evaluation can be accessed in the original development, internal validation manuscripts and implementation manuscripts.^18,28,32^

### Evaluation

To answer our first research objective, we evaluated the model performance in the new healthcare setting using precision, recall, area under the precision-recall curve (AUPRC), and area under the receiver operator curve (AUROC). To better understand the performance of SepsisWatch across the different ED and hospital sites, we separately evaluated the model at each location within Summa Health.

To answer our second research objective, we conduct several additional analyses. First, to estimate the potential effect of model integration on clinical care, we quantified the average ‘lead time’, defined as the amount of time between a ‘high risk’ model prediction and a patient meeting sepsis criterion. This measure helps quantify the potential opportunity for earlier intervention. Lead time is only measured for true positive cases identified early by SepsisWatch. Second, to assess the workflow burden placed on staff, we assessed the number of alerts that would be sent to either a charge nurse or rapid response team (RRT) at a given model threshold. The lead time and number of alerts are calculated separately for each Summa Health site.

## Results

### Cohort Characteristics

In total 205,005 encounters from 101,584 unique patients met inclusion criteria for the study. Most patients were female (54.7%, n = 112,223) and the mean age was 50 (IQR, [38,71]). The incidence of sepsis within the first 36 hours of encounters was 3.38% (n = 6,920). The majority of encounters were initiated at the two EDs co-located with acute care hospitals: ACH Emergency Department (ED) (59.08%, n=121,131) and SHB ED (23.53%, n = 48,244). The remaining encounters were initiated at the two standalone EDs: ACH Green ED (11.11%, n = 22,893) and SHB Wadsworth ED (6.21%, n = 12,737). Patient characteristics are broken down by site in **Table 1**.

**Table 1:** Description of cohort characteristics across all four emergency departments in the Summa Health system.

| **Baseline Characteristics of Cohort** | **ACH EMERGENCY DEPT** | **SHB EMERGENCY DEPT** | **ACH GREEN ED** | **SHB WADSWORTH ED** |
| --- | --- | --- | --- | --- |
| **Total Encounters, N (%)** | **121131 (59.08%)** | **48244 (23.53%)** | **22893 (11.11%)** | **12737 (6.21%)** |
| **Age (years), mean ±SD** | 50.92 ± 20.12 | 50.71 ± 20.18 | 47.36 ± 19.28 | 48 ± 20.05 |
| **Sex Male, N (%)** | 56540 (46.67%) | 21,284 (44.12%) | 9,552 (41.72%) | 5,551 (43.58%) |
| **Sex Female, N (%)** | 64470 (53.22%) | 26,960 (55.88%) | 13,341 (58.28%) | 7,184 (56.40%) |
| **Sex (Others/Missing), N (%)** | 121 (0.10%) | 0 (0%) | 0 (0%) | 2 (0.02%) |
| **Race, N (%)** |  |  |  |  |
| Black or African American | 36431 (30.08%) | 6,545 (13.57%) | 2,436 (10.64%) | 279 (2.19%) |
| Caucasian/White | 75833 (62.60%) | 40,754 (84.47%) | 20,086 (87.74%) | 12,242 (96.11%) |
| Missing/other | 8867 (7.32%) | 945 (1.96%) | 371 (1.62%) | 216 (1.70%) |
| **Comorbidities, N (%)** |  |  |  |  |
| Congestive heart failure | 4515 (3.73%) | 1,222 (2.53%) | 257 (1.12%) | 192 (1.51%) |
| Hypertension | 5805 (4.80%) | 1,581 (3.28%) | 534 (2.33%) | 479 (3.80%) |
| Pulmonary circulation disorders | 5441 (4.49%) | 2,777 (5.76%) | 633 (2.77%) | 459 (3.60%) |
| Diabetes mellitus | 3702 (3.06%) | 1,012 (2.10%) | 283 (1.24%) | 205 (1.61%) |
| Fluid and electrolyte disorders | 9324 (7.70%) | 3,705 (7.68%) | 995 (4.35%) | 642 (5.04%) |
| Depression | 2521 (2.08%) | 612 (1.27%) | 141 (0.62%) | 161 (1.26%) |
| **In-hospital mortality, N (%)** | 1139 (0.94%) | 381 (0.79%) | 0.16% | 0.28% |
| **Median Length of Stay (25%–75%)** | 6.38 (3.5, 56.7) | 4 (2.46, 28.8) | 2.63 (1.71, 4.15) | 2.43 (1.6, 4.31) |
| **Overall rate of ICU admission, N (%)** | 5487 (4.53%) | 2644 (5.48%) | 272 (1.19%) | 320 (2.51%) |
| **Septic, N (%)** | 4639 (3.83%) | 1785 (3.70%) | 240 (1.05%) | 245 (1.92%) |

### Research Objective 1 - Model Performance

Overall, the SepsisWatch model demonstrated robust performance on the geographically and temporally distinct Summa Health patient population. Moreover, there was little variation in model performance across the different emergency departments. Area under the precision-recall curve (AUPRC) for the four sites was 0.252 for ACH ED, 0.248 for SHB ED, 0.177 for ACH Green ED and 0.216 for SHB Wadsworth ED. The area under the receiver operator curve (AUROC) for the four sites was 0.919 for ACH ED, 0.906 for SHB ED, 0.960 for ACH Green ED and 0.928 for SHB Wadsworth ED. The AUROC and AUPRC for the four sites are visualized in **Figure 1** and **Figure 2**. The model performed similarly at each distinct location when stratifying patient population by White and Black races (**Table 2**).

**Figure 1:**
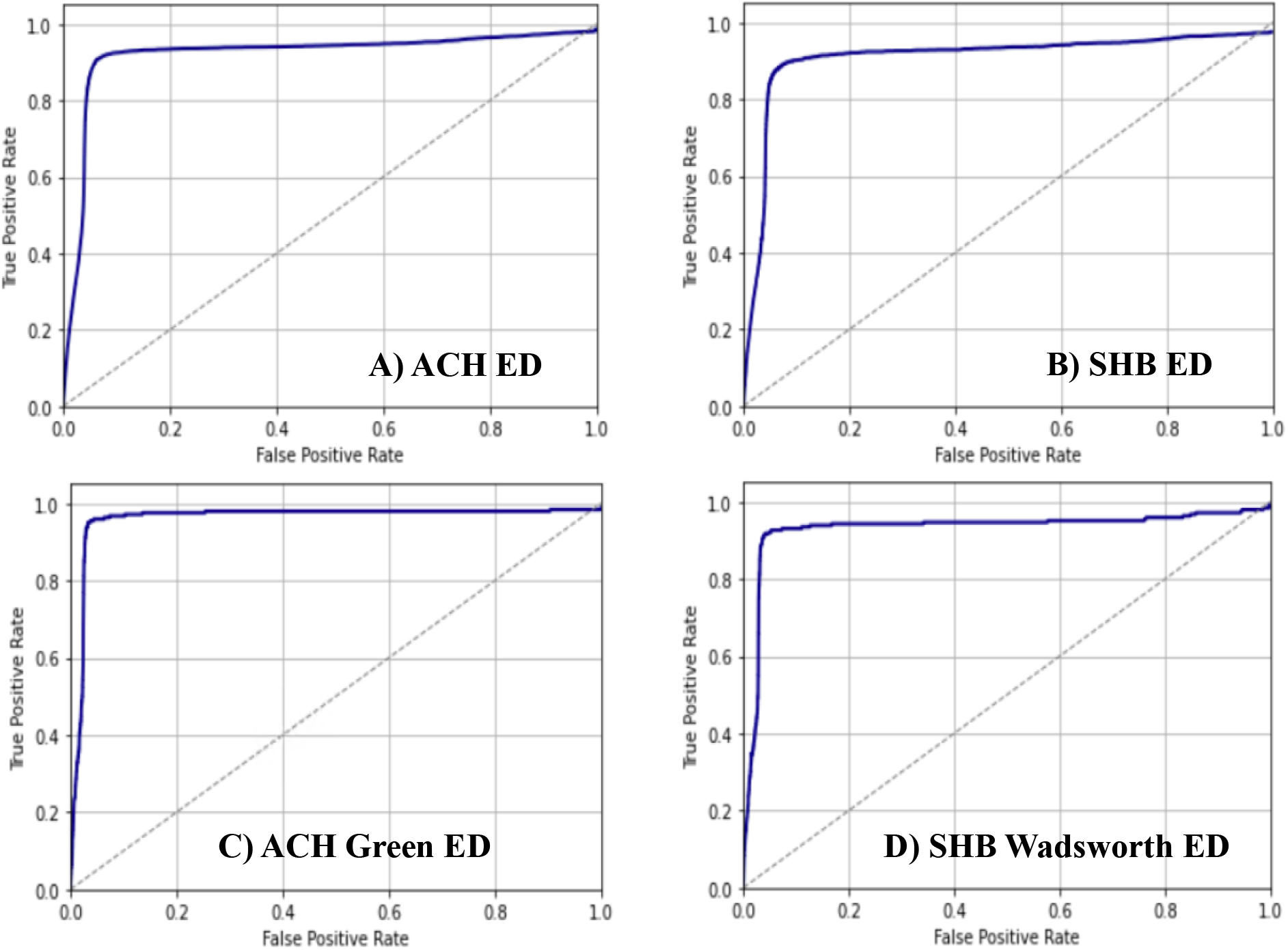
Area under the receiver operator curve at each of Summa Health’s four emergency departments, A) ACH Emergency Department (ED), B) SHB ED, C) ACH Green ED, D) SHB Wadsworth ED

**Figure 2:**
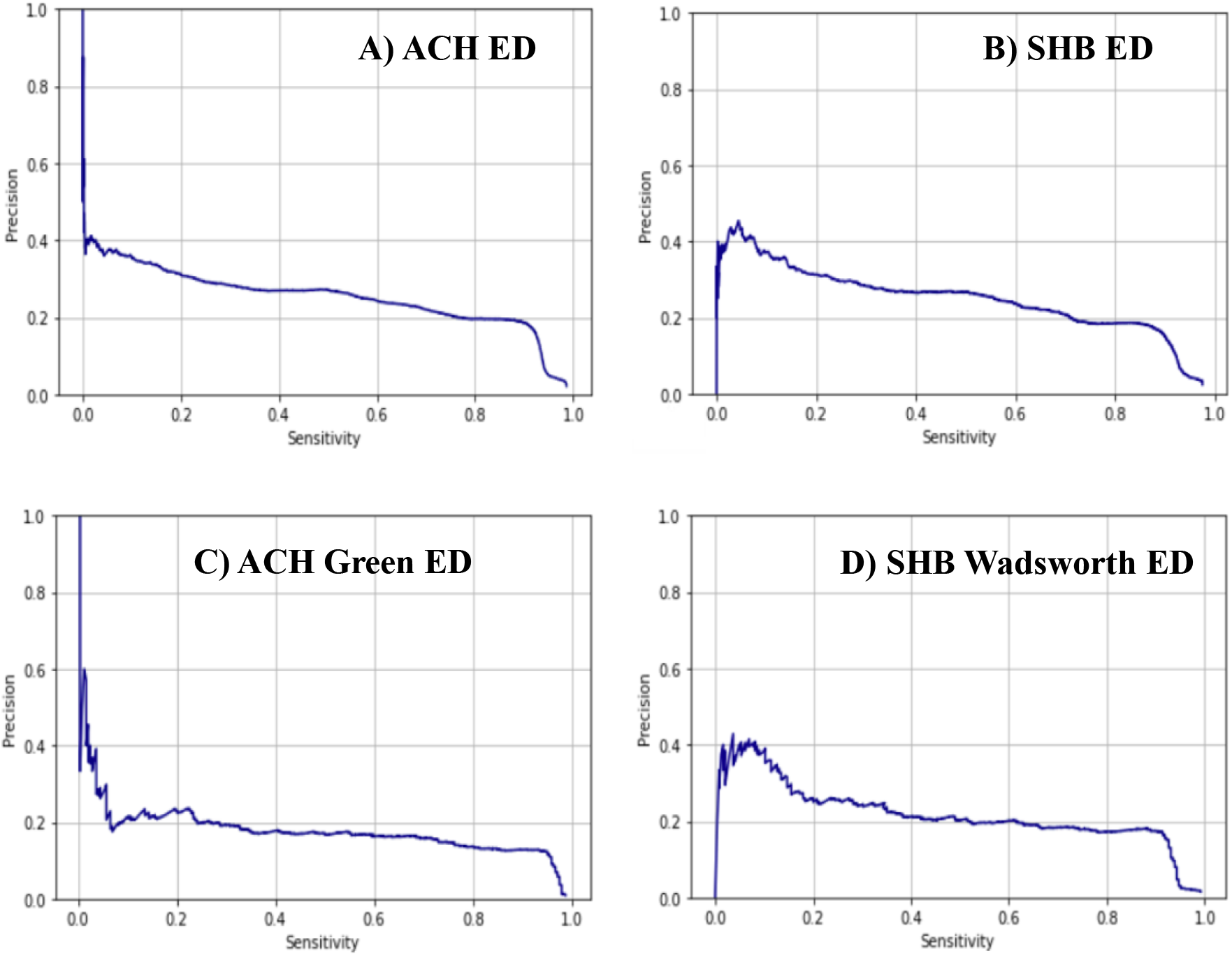
Area under precision-recall curve at each of Summa Health’s four emergency departments A) ACH Emergency Department (ED) B) SHB ED C) ACH Green ED D) SHB Wadsworth ED

**Table 2:** Site-specific performance metrics stratified by race including area under the precision recall curve (AU-PR) and area under the receiver operator curve (AUROC)

| Cohort | AU-PR | AUROC |
| --- | --- | --- |
| **ACH ED** |  |  |
| Entire Population | 0.252 | 0.919 |
| White Adults | 0.255 | 0.918 |
| Black Adults | 0.238 | 0.911 |
| **SHB ED** |  |  |
| Entire Population | 0.248 | 0.906 |
| White Adults | 0.246 | 0.901 |
| Black Adults | 0.286 | 0.920 |
| **ACH Green ED** |  |  |
| Entire Population | 0.177 | 0.960 |
| White Adults | 0.175 | 0.957 |
| Black Adults | 0.18 | 0.983 |
| **SHB- Wadsworth ED** |  |  |
| Entire Population | 0.216 | 0.928 |
| White Adults | 0.213 | 0.926 |
| Black Adults | 0.325 | 0.974 |

### Research Objective 2 – Evaluating Potential Clinical Benefit and Alert Fatigue

Model performance measures, including precision, recall, average number of alerts per day, and average lead time prior to meeting sepsis criteria vary based on model threshold. Performance measures across thresholds are illustrated in **Table 3**. If a threshold is set at each site to fix precision (positive predictive value) at 20%, the recall (sensitivity) is 76.2% at ACH ED, 70.9% at SHB ED, 27.0% at ACH Green ED, and 61.2% at SHB Wadsworth ED. At this same threshold, the average number of alerts per day is 7 at ACH ED, 3 at SHB ED, 2 at ACH Green ED, and 1 at SHB Wadsworth ED. Lastly, the average lead time (hours) is 4.06 at ACH ED, 3.79 at SHB ED, 5.07 at ACH Green ED, and 3.58 at SHB Wadsworth ED (**eTable 1 in the Supplement**).

**Table 3:**
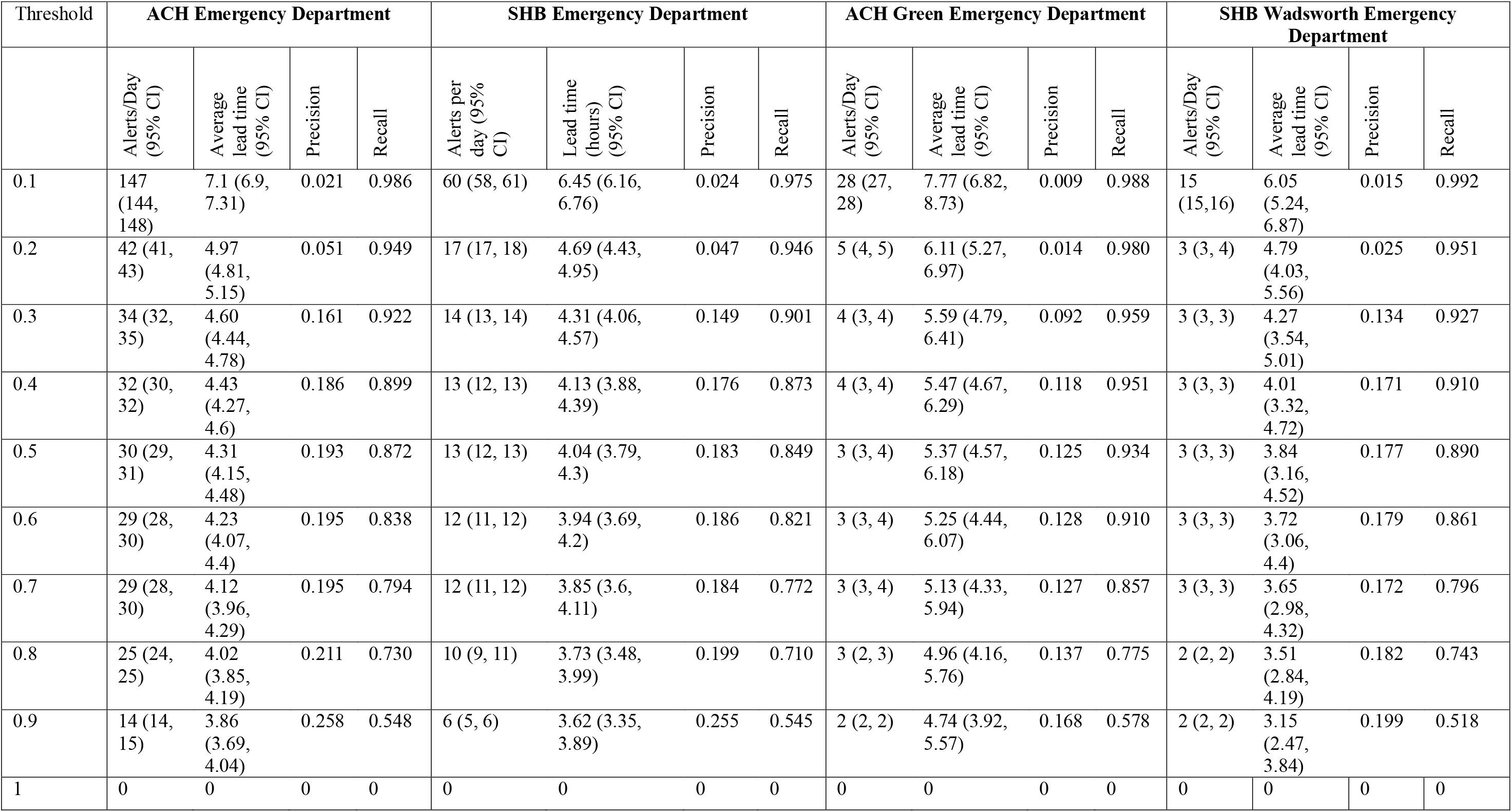
Site specific model performance including precision, recall, average number of alerts per day and average lead time based on model threshold.

| Threshold | ACH Emergency Department |  |  |  | SHB Emergency Department |  |  |  | ACH Green Emergency Department |  |  |  | SHB Wadsworth Emergency Department |  |  |  |
| --- | --- | --- | --- | --- | --- | --- | --- | --- | --- | --- | --- | --- | --- | --- | --- | --- |
|  | Alerts/Day<br>(95% CI) | Average<br>lead time<br>(95% CI) | Precision | Recall | Alerts per<br>day (95%<br>CI) | Lead time<br>(hours)<br>(95% CI) | Precision | Recall | Alerts/Day<br>(95% CI) | Average<br>lead time<br>(95% CI) | Precision | Recall | Alerts/Day<br>(95% CI) | Average<br>lead time<br>(95% CI) | Precision | Recall |
| 0.1 | 147<br>(144,<br>148) | 7.1 (6.9,<br>7.31) | 0.021 | 0.986 | 60 (58, 61) | 6.45 (6.16,<br>6.76) | 0.024 | 0.975 | 28 (27,<br>28) | 7.77 (6.82,<br>8.73) | 0.009 | 0.988 | 15<br>(15,16) | 6.05<br>(5.24,<br>6.87) | 0.015 | 0.992 |
| 0.2 | 42 (41,<br>43) | 4.97<br>(4.81,<br>5.15) | 0.051 | 0.949 | 17 (17, 18) | 4.69 (4.43,<br>4.95) | 0.047 | 0.946 | 5 (4, 5) | 6.11 (5.27,<br>6.97) | 0.014 | 0.980 | 3 (3, 4) | 4.79<br>(4.03,<br>5.56) | 0.025 | 0.951 |
| 0.3 | 34 (32,<br>35) | 4.60<br>(4.44,<br>4.78) | 0.161 | 0.922 | 14 (13, 14) | 4.31 (4.06,<br>4.57) | 0.149 | 0.901 | 4 (3, 4) | 5.59 (4.79,<br>6.41) | 0.092 | 0.959 | 3 (3, 3) | 4.27<br>(3.54,<br>5.01) | 0.134 | 0.927 |
| 0.4 | 32 (30,<br>32) | 4.43<br>(4.27,<br>4.6) | 0.186 | 0.899 | 13 (12, 13) | 4.13 (3.88,<br>4.39) | 0.176 | 0.873 | 4 (3, 4) | 5.47 (4.67,<br>6.29) | 0.118 | 0.951 | 3 (3, 3) | 4.01<br>(3.32,<br>4.72) | 0.171 | 0.910 |
| 0.5 | 30 (29,<br>31) | 4.31<br>(4.15,<br>4.48) | 0.193 | 0.872 | 13 (12, 13) | 4.04 (3.79,<br>4.3) | 0.183 | 0.849 | 3 (3, 4) | 5.37 (4.57,<br>6.18) | 0.125 | 0.934 | 3 (3, 3) | 3.84<br>(3.16,<br>4.52) | 0.177 | 0.890 |
| 0.6 | 29 (28,<br>30) | 4.23<br>(4.07,<br>4.4) | 0.195 | 0.838 | 12 (11, 12) | 3.94 (3.69,<br>4.2) | 0.186 | 0.821 | 3 (3, 4) | 5.25 (4.44,<br>6.07) | 0.128 | 0.910 | 3 (3, 3) | 3.72<br>(3.06,<br>4.4) | 0.179 | 0.861 |
| 0.7 | 29 (28,<br>30) | 4.12<br>(3.96,<br>4.29) | 0.195 | 0.794 | 12 (11, 12) | 3.85 (3.6,<br>4.11) | 0.184 | 0.772 | 3 (3, 4) | 5.13 (4.33,<br>5.94) | 0.127 | 0.857 | 3 (3, 3) | 3.65<br>(2.98,<br>4.32) | 0.172 | 0.796 |
| 0.8 | 25 (24,<br>25) | 4.02<br>(3.85,<br>4.19) | 0.211 | 0.730 | 10 (9, 11) | 3.73 (3.48,<br>3.99) | 0.199 | 0.710 | 3 (2, 3) | 4.96 (4.16,<br>5.76) | 0.137 | 0.775 | 2 (2, 2) | 3.51<br>(2.84,<br>4.19) | 0.182 | 0.743 |
| 0.9 | 14 (14,<br>15) | 3.86<br>(3.69,<br>4.04) | 0.258 | 0.548 | 6 (5, 6) | 3.62 (3.35,<br>3.89) | 0.255 | 0.545 | 2 (2, 2) | 4.74 (3.92,<br>5.57) | 0.168 | 0.578 | 2 (2, 2) | 3.15<br>(2.47,<br>3.84) | 0.199 | 0.518 |
| 1 | 0 | 0 | 0 | 0 | 0 | 0 | 0 | 0 | 0 | 0 | 0 | 0 | 0 | 0 | 0 | 0 |

Alternatively, if a threshold is set at each site to fix recall (sensitivity) at 60%, the precision (positive predictive value) is 23.9% at ACH ED, 23.1% at SHB ED, 16.3% at ACH Green ED, and 20.1% at SHB Wadsworth ED. At this same threshold, the average number of alerts per day is 7 at ACH ED, 3 at SHB ED, 1 at ACH Green ED, and 1 at SHB Wadsworth ED. Lastly, the average lead time (hours) is 3.94 at ACH ED, 3.69 at SHB ED, 4.89 at ACH Green ED, and 3.42 at SHB Wadsworth ED (**eTable 2 in the Supplement**).

## Discussion

In this study we present the first external validation of a sepsis ML model in a community based health setting. The model was originally developed at a tertiary academic medical center in North Carolina and maintained robust performance across four EDs at a community health system in Ohio. The original model achieved an AUROC of 0.83 and an AUPRC of 0.257.^18^ This strong performance was maintained in the external validation presented in this study, in which SepsisWatch achieved an AUROC of 0.92 and an AUPRC of 0.24. Moreover, SepsisWatch performed strongly across two EDs that are co-located with acute care hospitals and two standalone EDs, marking the first known validation of a sepsis machine learning algorithm in a standalone ED. A ‘Model Facts’ sheet containing details of this external validation and the original model development can be seen in **eFigure 1 in the Supplement.**

Most machine learning algorithms are not externally validated and those that are demonstrate varied performance. Wong ***et al.*** demonstrated that the proprietary Epic Sepsis Model (ESM) had substantially worse calibration and discrimination among adult patients within one US academic health system. A later study by Lyons ***et al.*** also revealed and varied performance of ESM across nine hospitals and found that hospitals with lower sepsis incidence had worse AUROC.^24^ On the other hand, Brajer ***et al.*** demonstrated robust performance of an in-hospital mortality ML model across multiple hospitals within the same health system.^25^ Moor *et al.* used a novel validation technique pooling predictions of models developed on different data, achieving an AUROC of 0.76 on external validation cohorts verse 0.84 on internal validation.^33^ While Moor *et al.* were able improve external validation performance after fine tuning the models using a small set of data from the target testing site, performance remained stronger in the internal validation cohort.^33^ The new version of the ESM similarly includes fine tuning on local data to improve performance, but improved performance of that model has not yet been reported in the peer-reviewed literature. In addition, local fine tuning necessitates additional expertise, personnel, and compute infrastructure.

Beyond generalizing across time and geographic settings, SepsisWatch also exhibited strong and robust performance across demographic subgroups. Unfortunately, examination of ML model performance across racial subgroups has not been reported in many prior external validation studies in sepsis and other clinical domains.^27,33^ There are significant concerns when models are trained on datasets with minimal diversity and tested on populations that include more representation of historically marginalized populations.^34^ The current study builds on prior work from dermatology that found that training on more diverse datasets led to improved performance on diverse populations.^35^ SepsisWatch was trained on a cohort of adults in North Carolina that were 30% Black and performed well on cohorts of adults in Ohio that were 3% – 30% Black. The robust performance of the SepsisWatch model presented in the current study emphasizes the importance of training models on diverse datasets.

Balancing alert fatigue with clinically meaningful predictions is a well-established challenge in operationalizing sepsis prediction models.^27,36,37^ Moreover, given expected trade-offs between model precision and recall, choosing a prediction threshold that balances both overall performance with clinical impact becomes exceedingly important. For example, at the ACH Emergency Department, when prediction threshold was set to 0.1, the model would send on average 147 alerts per day. At this threshold the model would identify almost every instance of sepsis and provide an average lead time of 7.1 hours. But the precision of the model at this threshold is 2.1% and approximately 47 alerts need to be screened for every case that goes on to develop sepsis. In contrast, a prediction threshold of 0.6 would afford a recall of 84%, precision of 20% and average lead time of 4.23 hours. The number of alerts at this threshold is reduced 80% to an average of 29 alerts per day. Thus, operational leaders within each setting can titrate the model threshold to align with the capacity of front-line clinicians to respond to alerts. Ultimately, the threshold should be determined after a silent trial in which front-line clinicians can help test SepsisWatch and ensure that patients flagged by the model are appropriate for review.

Our study had several limitations. First, this was a retrospective study and while model performance and potential opportunity to improve care could be assessed, SepsisWatch will need to be prospectively tested to measure impact. Second, this study features a single health system in a single geography. The generalizability of SepsisWatch beyond North Carolina and Ohio remains unknown. Third, this study did not directly compare SepsisWatch to other available sepsis models, such as the Epic Sepsis Model or TREWS. Those technologies were not available to include in the analysis and additional contracting and potential costs would be required to compare multiple algorithms. Future comparative effectiveness research will be required to better understand the tradeoffs associated with use of each model. Fourth, this study evaluated SepsisWatch during a time window that included the COVID-19 pandemic. The time window was two years and covered multiple variant waves of the virus. Future research will be needed to characterize variability of model performance during viral outbreaks. Finally, this study only evaluated the performance of SepsisWatch with an 8-hour snooze window. Future studies are necessary to optimize and tailor this time window for different implementation sites.

## Conclusions

This study illustrates the first successful external validation of a sepsis deep learning model, SepsisWatch, within a community health system. SepsisWatch demonstrated remarkable consistency in performance despite variations in time, geography, and patient demographics. The strong performance of SepsisWatch highlights the possibility of effectively transporting advanced ML models from academic to community based hospital settings. Additionally, the study addresses the crucial balance between minimizing alert fatigue and maintaining clinical relevance, emphasizing the importance of carefully selecting prediction thresholds tailored to each deployment site and clinical context. The findings suggest that while there are challenges in creating more broadly applicable clinical prediction models, careful evaluation in different health contexts is feasible and can yield promising results. This study will pave the way for future prospective studies to measure the clinical and operational effect of SepsisWatch and other sepsis machine learning models integrated into clinical care.

## Data Availability

All data produced in the present study are available upon reasonable request to the authors

## References

1. Levy MM, Fink MP, Marshall JC, et al. 2001 SCCM/ESICM/ACCP/ATS/SIS International Sepsis Definitions Conference. Intensive Care Med. 2003;29(4):530–538. doi:10.1007/s00134-003-1662-x

2. Liu V, Escobar GJ, Greene JD, et al. Hospital deaths in patients with sepsis from 2 independent cohorts. JAMA. 2014;312(1):90–92. doi:10.1001/jama.2014.5804

3. Rudd KE, Johnson SC, Agesa KM, et al. Global, regional, and national sepsis incidence and mortality, 1990-2017: analysis for the Global Burden of Disease Study. Lancet Lond Engl. 2020;395(10219):200–211. doi:10.1016/S0140-6736(19)32989-7

4. Buchman TG, Simpson SQ, Sciarretta KL, et al. Sepsis Among Medicare Beneficiaries: 1. The Burdens of Sepsis, 2012-2018. Crit Care Med. 2020;48(3):276–288. doi:10.1097/CCM.0000000000004224

5. Paoli CJ, Reynolds MA, Sinha M, Gitlin M, Crouser E. Epidemiology and Costs of Sepsis in the United States-An Analysis Based on Timing of Diagnosis and Severity Level. Crit Care Med. 2018;46(12):1889–1897. doi:10.1097/CCM.0000000000003342

6. Rhodes A, Evans LE, Alhazzani W, et al. Surviving Sepsis Campaign: International Guidelines for Management of Sepsis and Septic Shock: 2016. Intensive Care Med. 2017;43(3):304–377. doi:10.1007/s00134-017-4683-6

7. Cohen J, Vincent JL, Adhikari NKJ, et al. Sepsis: a roadmap for future research. Lancet Infect Dis. 2015;15(5):581–614. doi:10.1016/S1473-3099(15)70112-X

8. Kumar A, Roberts D, Wood KE, et al. Duration of hypotension before initiation of effective antimicrobial therapy is the critical determinant of survival in human septic shock. Crit Care Med. 2006;34(6):1589–1596. doi:10.1097/01.CCM.0000217961.75225.E9

9. Ferrer R, Martin-Loeches I, Phillips G, et al. Empiric antibiotic treatment reduces mortality in severe sepsis and septic shock from the first hour: results from a guideline-based performance improvement program. Crit Care Med. 2014;42(8):1749–1755. doi:10.1097/CCM.0000000000000330

10. Liu VX, Fielding-Singh V, Greene JD, et al. The Timing of Early Antibiotics and Hospital Mortality in Sepsis. Am J Respir Crit Care Med. 2017;196(7):856–863. doi:10.1164/rccm.201609-1848OC

11. Peltan ID, Brown SM, Bledsoe JR, et al. ED Door-to-Antibiotic Time and Long-term Mortality in Sepsis. Chest. 2019;155(5):938–946. doi:10.1016/j.chest.2019.02.008

12. Rhee C, Yu T, Wang R, et al. Association Between Implementation of the Severe Sepsis and Septic Shock Early Management Bundle Performance Measure and Outcomes in Patients With Suspected Sepsis in US Hospitals. JAMA Netw Open. 2021;4(12):e2138596. doi:10.1001/jamanetworkopen.2021.38596

13. Townsend SR, Phillips GS, Duseja R, et al. Effects of Compliance With the Early Management Bundle (SEP-1) on Mortality Changes Among Medicare Beneficiaries With Sepsis: A Propensity Score Matched Cohort Study. Chest. 2022;161(2):392–406. doi:10.1016/j.chest.2021.07.2167

14. Barbash IJ, Davis BS, Yabes JG, Seymour CW, Angus DC, Kahn JM. Treatment Patterns and Clinical Outcomes After the Introduction of the Medicare Sepsis Performance Measure (SEP-1). Ann Intern Med. 2021;174(7):927–935. doi:10.7326/M20-5043

15. Henry KE, Hager DN, Pronovost PJ, Saria S. A targeted real-time early warning score (TREWScore) for septic shock. Sci Transl Med. 2015;7(299):299ra122. doi:10.1126/scitranslmed.aab3719

16. Desautels T, Calvert J, Hoffman J, et al. Prediction of Sepsis in the Intensive Care Unit With Minimal Electronic Health Record Data: A Machine Learning Approach. JMIR Med Inform. 2016;4(3):e28. doi:10.2196/medinform.5909

17. Horng S, Sontag DA, Halpern Y, Jernite Y, Shapiro NI, Nathanson LA. Creating an automated trigger for sepsis clinical decision support at emergency department triage using machine learning. PLOS ONE. 2017;12(4):e0174708. doi:10.1371/journal.pone.0174708

18. Bedoya AD, Futoma J, Clement ME, et al. Machine learning for early detection of sepsis: an internal and temporal validation study. JAMIA Open. 2020;3(2):252–260. doi:10.1093/jamiaopen/ooaa006

19. Tarabichi Y, Cheng A, Bar-Shain D, et al. Improving Timeliness of Antibiotic Administration Using a Provider and Pharmacist Facing Sepsis Early Warning System in the Emergency Department Setting: A Randomized Controlled Quality Improvement Initiative. Crit Care Med. 2022;50(3):418–427. doi:10.1097/CCM.0000000000005267

20. Adams R, Henry KE, Sridharan A, et al. Prospective, multi-site study of patient outcomes after implementation of the TREWS machine learning-based early warning system for sepsis. Nat Med. 2022;28(7):1455–1460. doi:10.1038/s41591-022-01894-0

21. Shimabukuro DW, Barton CW, Feldman MD, Mataraso SJ, Das R. Effect of a machine learning-based severe sepsis prediction algorithm on patient survival and hospital length of stay: a randomised clinical trial. BMJ Open Respir Res. 2017;4(1):e000234. doi:10.1136/bmjresp-2017-000234

22. Nelson JL, Smith BL, Jared JD, Younger JG. Prospective trial of real-time electronic surveillance to expedite early care of severe sepsis. Ann Emerg Med. 2011;57(5):500–504. doi:10.1016/j.annemergmed.2010.12.008

23. Giannini HM, Ginestra JC, Chivers C, et al. A Machine Learning Algorithm to Predict Severe Sepsis and Septic Shock: Development, Implementation, and Impact on Clinical Practice. Crit Care Med. 2019;47(11):1485–1492. doi:10.1097/CCM.0000000000003891

24. Lyons PG, Hofford MR, Yu SC, et al. Factors Associated With Variability in the Performance of a Proprietary Sepsis Prediction Model Across 9 Networked Hospitals in the US. JAMA Intern Med. 2023;183(6):611–612. doi:10.1001/jamainternmed.2022.7182

25. Brajer N, Cozzi B, Gao M, et al. Prospective and External Evaluation of a Machine Learning Model to Predict In-Hospital Mortality of Adults at Time of Admission. JAMA Netw Open. 2020;3(2):e1920733. doi:10.1001/jamanetworkopen.2019.20733

26. Bedoya AD, Clement ME, Phelan M, Steorts RC, O’Brien C, Goldstein BA. Minimal Impact of Implemented Early Warning Score and Best Practice Alert for Patient Deterioration. Crit Care Med. 2019;47(1):49–55. doi:10.1097/CCM.0000000000003439

27. Wong A, Otles E, Donnelly JP, et al. External Validation of a Widely Implemented Proprietary Sepsis Prediction Model in Hospitalized Patients. JAMA Intern Med. 2021;181(8):1065–1070. doi:10.1001/jamainternmed.2021.2626

28. Sendak MP, Ratliff W, Sarro D, et al. Real-World Integration of a Sepsis Deep Learning Technology Into Routine Clinical Care: Implementation Study. JMIR Med Inform. 2020;8(7):e15182. doi:10.2196/15182

29. Sendak M, Sirdeshmukh G, Ochoa T, et al. Development and Validation of ML-DQA -- a Machine Learning Data Quality Assurance Framework for Healthcare. Published online August 4, 2022. doi:10.48550/arXiv.2208.02670

30. Topiwala R, Patel K, Twigg J, Rhule J, Meisenberg B. Retrospective Observational Study of the Clinical Performance Characteristics of a Machine Learning Approach to Early Sepsis Identification. Crit Care Explor. 2019;1(9):e0046. doi:10.1097/CCE.0000000000000046

31. Ginestra JC, Giannini HM, Schweickert WD, et al. Clinician Perception of a Machine Learning-Based Early Warning System Designed to Predict Severe Sepsis and Septic Shock. Crit Care Med. 2019;47(11):1477–1484. doi:10.1097/CCM.0000000000003803

32. Futoma J, Hariharan S, Heller K, et al. An Improved Multi-Output Gaussian Process RNN with Real-Time Validation for Early Sepsis Detection. In: Proceedings of the 2nd Machine Learning for Healthcare Conference. PMLR; 2017:243–254. Accessed September 26, 2023. https://proceedings.mlr.press/v68/futoma17a.html

33. Moor M, Bennett N, Plečko D, et al. Predicting sepsis using deep learning across international sites: a retrospective development and validation study. eClinicalMedicine. 2023;62:102124. doi:10.1016/j.eclinm.2023.102124

34. Chen IY, Pierson E, Rose S, Joshi S, Ferryman K, Ghassemi M. Ethical Machine Learning in Healthcare. Annu Rev Biomed Data Sci. 2021;4:123–144. doi:10.1146/annurev-biodatasci-092820-114757

35. Daneshjou R, Vodrahalli K, Novoa RA, et al. Disparities in dermatology AI performance on a diverse, curated clinical image set. Sci Adv. 2022;8(32):eabq6147. doi:10.1126/sciadv.abq6147

36. Gregory ME, Russo E, Singh H. Electronic Health Record Alert-Related Workload as a Predictor of Burnout in Primary Care Providers. Appl Clin Inform. 2017;08(3):686–697. doi:10.4338/ACI-2017-01-RA-0003

37. Borowski M, Görges M, Fried R, Such O, Wrede C, Imhoff M. Medical device alarms. 2011;56(2):73–83. doi:10.1515/bmt.2011.005

