## Supplement for "Evaluating the Portability of SepsisWatch: A Multi-Site External Validation of a Sepsis Machine Learning Model"

**Supplemental Material**

| **Emergency Department (ED)** | **PPV (%)** | **Recall (%)** | **Alerts/Day (95% CI)** | **Average Lead Time (hours)** |
|---|---|---|---|---|
| ACH ED | 20 | 76.2 | 28 (27.0, 29.0) | 4.06 (3.9, 4.3) |
| SHB ED | 20 | 70.9 | 11 (10.0,11.0) | 3.79 (3.2, 5.5) |
| ACH Green ED | 20 | 27.0 | 1 (1.0, 1.0) | 5.07 (3.2, 5.6) |
| SHB Wadsworth ED | 20 | 61.2 | 2 (2.0, 2.0) | 3.58 (3.4, 3.9) |

**eTable 1:** Site specific model performance at a fixed positive predictive value (PPV) of 20% including recall, average number of alerts per day and average lead time

| **Emergency Department (ED)** | **Recall (%)** | **PPV (%)** | **Alerts/Day** | **Average Lead Time (hours)** |
|---|---|---|---|---|
| ACH ED | 60 | 23.9 | 19 (18, 19) | 3.94 (3.77, 4.11) |
| SHB ED | 60 | 23.1 | 7 (7.0, 8.0) | 3.69 (3.42, 3.95) |
| ACH Green ED | 60 | 16.3 | 2 (2.0, 2.0) | 4.89 (4.0, 5.63) |
| SHB Wadsworth ED | 60 | 20.1 | 2 (2.0, 2.0) | 3.42 (2.72, 4.11) |

**eTable 2:** Site specific model performance at a fixed recall of 60% including positive predictive value (PPV), average number of alerts per day and average lead time

### *Model Facts*

**Model name:** Deep Sepsis

**Locale:** Summa Health

**Approval Date:** 09/22/2019 **Last Update: 11/13/2023** **Version:** 1.0


#### Summary

This model uses EHR input data collected from a patient's current inpatient encounter to estimate the probability that the patient will meet sepsis criteria within the next 4 hours. It was developed in 2016-2019 by the Duke Institute for Health Innovation. The model was licensed to Cohere Med in July 2019.


#### Mechanism

- **Outcome** ……………………………………………sepsis within the next 4 hours, see outcome definition in "Other Information"
- **Output** ……………………………………………………………0% - 100% probability of sepsis occurring in the next 4 hours
- **Target population** ……………………………………………all adult patients >18 y.o. presenting to DUH ED
- **Time of prediction** ……………………………………………every hour of a patient's encounter
- **Input data source**……………………………………………electronic health record (EHR)
- **Input data type** ……………………………………………demographics, analytes, vitals, medication administrations
- **Training data location and time-period** ……………………DUH, diagnostic cohort, 10/2014 – 12/2015
- **Model type**…………………………………………………… Recurrent Neural Network

#### Validation and performance

| | Prevalence | AUC | PPV @ Sensitivity of 60% | Sensitivity @ PPV of 20% | Cohort Type | Cohort URL / DOI |
|---|---|---|---|---|---|---|
| **Local Retrospective** | 18.9% | 0.88 | 0.14 | 0.50 | Diagnostic | arxiv.org/abs/1708.05894 |
| **Local Temporal** | 6.4% | 0.94 | 0.20 | 0.66 | Diagnostic | jmir.org/preprint/15182 |
| **Local Prospective** | **TBD** | **TBD** | **TBD** | **TBD** | **TBD** | **TBD** |
| **External** | 3.38% | 0.92 | 0.235 | 0.735 | Diagnostic | **TBD** |
| **Target Population** | 6.4% | 0.94 | 0.20 | 0.66 | Diagnostic | jmir.org/preprint/15182 |

#### Uses and directions

- **Benefits:** Early identification and prompt treatment of sepsis can improve patient morbidity and mortality.
- **Target population and use case:** Every hour, data is pulled from the EHR to calculate risk of sepsis for every patient at the DUH ED. A rapid response team nurse reviews every high-risk patient with a physician in the ED to confirm whether or not to initiate treatment for sepsis.
- **General use:** This model is intended to be used to by clinicians to identify patients for further assessment for sepsis. The model is not a diagnostic for sepsis and is not meant to guide or drive clinical care. This model is intended to complement other pieces of patient information related to sepsis as well as a physical evaluation to determine the need for sepsis treatment.
- **Appropriate decision support:** The model identifies patient X as at a high risk of sepsis. A rapid response team nurse discusses the patient with the ED physician caring for the patient and they agree the patient does not require treatment for sepsis.
- **Before using this model:** Test the model retrospectively and prospectively on a diagnostic cohort that reflects the target population that the model will be used upon to confirm validity of the model within a local setting.
- **Safety and efficacy evaluation:** Analysis of data from clinical trial (NCT03655626) is underway. Preliminary data shows rapid response team, nurse-driven workflow was effective at improving sepsis treatment bundle compliance.

### Warnings

- **Risks:** Even if used appropriately, clinicians using this model can misdiagnose sepsis. Delays in a sepsis diagnosis can lead to morbidity and mortality. Patients who are incorrectly treated for sepsis can be exposed to risks associated with unnecessary antibiotics and intravenous fluids.
- **Inappropriate Settings:** This model was not trained or evaluated on patients receiving care in the ICU. Do not use this model in the ICU setting without further evaluation. This model was trained to identify the first episode of sepsis during an inpatient encounter. Do not use this model after an initial sepsis episode without further evaluation.
- **Clinical Rationale:** The model is not interpretable and does not provide rationale for high risk scores. Clinical end users are expected to place model output in context with other clinical information to make final determination of diagnosis.
- **Inappropriate decision support:** This model may not be accurate outside of the target population, primarily adults in the non-ICU setting. This model is not a diagnostic and is not designed to guide clinical diagnosis and treatment for sepsis.
- **Generalizability:** This model was primarily evaluated within the local setting of Duke University Hospital. Do not use this model in an external setting without further evaluation.
- **Discontinue use if:** Clinical staff raise concerns about utility of the model for the indicated use case or large, systematic changes occur at the data level that necessitates re-training of the model.

### Other information:

- **Outcome Definition:** https://doi.org/10.1101/648907
- **Related model:** http://doi.org/10.1001/jama.2016.0288
- **Model development & validation:** arxiv.org/abs/1708.05894
- **Model implementation**: jmir.org/preprint/15182
- **Clinical trial:** clinicaltrials.gov/ct2/show/NCT03655626
- **Clinical impact evaluation:** TBD
- **For inquiries and additional information:** please

**eFigure 1:** Updated 'Model Facts' sheet summarizing key information regarding model performance, validation, use context and warnings.
